# Impact of ‘EVEN FASTER’ concept to accelerate cervical cancer elimination in Norway: A model-based analysis

**DOI:** 10.1101/2023.11.06.23298170

**Authors:** Allison Portnoy, Kine Pedersen, Jane J. Kim, Emily A. Burger

## Abstract

**Background:** Experts have proposed an ‘EVEN FASTER’ concept targeting age-groups maintaining circulation of human papillomavirus (HPV). We explored the effects of these proposals compared with cervical cancer (CC) screening-based interventions on age-standardized incidence rate (ASR) and CC elimination (<4 cases per 100,000 women) timing in Norway.

**Methods:** We used a model-based approach to evaluate alternative HPV vaccination and CC screening scenarios compared with a status-quo scenario reflecting previous vaccination and screening. For cohorts ages 25–35 years, we examined 11 vaccination scenarios that incrementally increased vaccination coverage from current cohort-specific rates. Each vaccination scenario was coupled with three alternative screening strategies that varied the frequency of HPV-based screening. Population- and cohort-level outcomes included ASR, lifetime risk of CC, and colposcopy referrals.

**Results:** Several vaccination strategies coupled with de-intensified screening frequencies lowered population ASR, but did not accelerate CC elimination unless incremental vaccination coverage reached ∼90% for vaccine-naïve cohorts. Alternative strategies that increased screening adherence could both accelerate elimination and improve ASR compared to status-quo.

**Conclusions:** An ‘EVEN FASTER’ campaign is unlikely to accelerate CC elimination but may reduce population-level ASR. Alternatively, targeting under- and never-screeners may both eliminate CC faster and lead to greater health benefits compared with vaccination-based interventions.

## Background

Following the proposal of ‘HPV FASTER’ to extend eligible vaccination age and pair with screening tests,^1^ experts have recently proposed an ‘EVEN FASTER’ concept.^2^ The concept involves intensifying concomitant screening and vaccination campaigns to age-groups maintaining circulation of human papillomavirus (HPV) infection, the causal agent of nearly all cervical cancer. Advocates suggest this approach could both accelerate elimination of cervical cancer as a public health problem (defined as an incidence rate of <4 cases per 100,000 women^3^) and reduce the need for screening.^1,2,4^ While projections of elimination given current prevention efforts include Australia in 2028, USA in 2039–2046, and Norway in 2039,^5–7^ the ‘EVEN FASTER’ concept implies that these timeframes could be accelerated.

The role of vaccination on the elimination timeframe is unclear based on current evidence given the relatively long dwell time from HPV infection to cervical cancer.^8^ Models have estimated that most causal infections for women occur prior to the age of 25 years;^9^ for the women who acquire their causal infection later in life, it will take at least 10–20 years from the time of prevention of their causal infection to the prevention of an invasive cancer. In a US-based cervical cancer elimination analysis, increasing vaccination coverage did not result in projections of an earlier elimination timeframe given the already-high levels of vaccination, although this analysis did not include vaccination of mid-adult women.^6^ In contrast, the US analysis showed that improving screening coverage and follow-up adherence, particularly among women who screen infrequently, were likely to accelerate cervical cancer elimination.^6^ The introduction and implementation of HPV self-sampling as a screening strategy could further increase the impact among women who infrequently screen.^10^

Furthermore, it remains unclear whether vaccination of mid-adult women could reduce the need for cervical cancer screening. Previous studies have shown that screening frequency may be reduced among vaccinated women, and will be necessary to maintain the harm-benefit balance and cost-effectiveness of screening, but these studies generally focused on girls vaccinated in adolescence rather than catch-up programs of older age cohorts.^11,12^ While vaccination paired with less intensive screening strategies could be an important way to reduce dependence on screening and related resources and harms, there are uncertainties around vaccine efficacy in older women as well as on latent infections, which may contribute to overestimations of vaccine impact.^13^

Importantly, when deciding to de-intensify screening among fully or partially vaccinated cohorts, decision-makers must decide what benchmark of effectiveness they are willing to accept. Simulation models have been used to project the age-standardized incidence rates (ASR) among vaccine-eligible birth cohorts under alternative screening frequencies required to maintain harm-benefits ratios and cost-effectiveness.^11,12^ As a common benchmark comparator is based on current levels of screening and vaccination practice, the ASR may appear to increase in order to maintain efficiency (i.e., a similar ratio of health gains to costs incurred).^7,12^ Alternatively, a fully unvaccinated cohort of women screened under status-quo recommendations (e.g., 5-yearly screening) could be used as benchmark of an acceptable level of disease risk. De-intensifying screening alongside expanding vaccination coverage per the ‘EVEN FASTER’ concept could improve efficiency and harm-benefits ratio, but may not improve health benefits compared to a stringent status-quo of high vaccination coverage with frequent screening. Therefore, it is unclear whether ASR achieved under current screening guidelines should remain the benchmark for measuring effectiveness, as this population-level metric could mask relevant outcomes at the cohort level, such as cervical cancer cases averted, colposcopies avoided, and change in the lifetime risk of cervical cancer.^14^

Implementation of programs based on HPV ‘EVEN FASTER’ are time-sensitive in countries that introduced vaccination following approval and licensure, and may not be a one-size-fits-all recommendation. For example, as of 2023, vaccination coverage is high in Norway and few cohorts under age 35 years remain unvaccinated (women aged 33–35 years and boys 18–35 years remain fully naïve to direct vaccine protection, although likely receive considerable herd immunity benefits). In a recent white paper, the Cancer Registry of Norway proposes a new catch-up vaccination program (including re-vaccination of all men and women vaccinated with the first generation vaccines) for men and women up to age 30 years (inter alia) as an approach to expedite cervical cancer elimination.^15^ However, recent and ongoing vaccine shortages warrant careful evaluation of intensifying vaccination programs, particularly in settings like Norway with high coverage, prior to using resources that may not derive sufficient benefits.^16^

The HPV ‘EVEN FASTER’ hypotheses (acceleration of cervical cancer and reduction in the need for screening) need to be confirmed, first in terms of health impact and second in terms of efficiency. As a first step, we explored the health effects of the ‘EVEN FASTER’ proposals and alternative screening-based scenarios under context-specific contact patterns in Norway on cervical cancer burden and timing of cervical cancer elimination.

## Methods

### Scenarios and model assumptions

Using a multi-modeling approach that captured HPV transmission and cervical carcinogenesis,^5^ we estimated the ASRs, HPV prevalence rates, and lifetime cervical cancer risk associated with alternative 2-valent vaccination and screening scenarios compared with a status-quo scenario reflecting previous vaccination and screening for cohorts ages 25–35 years (including women previously targeted by temporary catch-up vaccination campaigns). In our base case, we examined 11 vaccination scenarios that incrementally increased vaccination coverage by a maximum of 20%, i.e., from 2021 cohort-specific rates for previously vaccinated cohorts to 90% for girls and 89% for boys and to 20% for vaccine-naïve female cohorts aged 33–35 (Figure 1) and vaccine-naïve male cohorts aged 18–35 (Figure S1) in 2023.^17^ These vaccination scenarios reflect the historic timing and age-specific receipt of any vaccine type relevant for prior routine and catch-up vaccination coverage of the specified cohorts. Norway introduced their temporary female catch-up program in 2016, seven years after their routine female 12-year-old program was implemented in 2009, and also switched from an initial 4-valent to 2-valent HPV vaccine program.^5,18^ A routine program for 12-year-old boys was introduced in 2018. Our base case assumed vaccine efficacy of 100% against HPV-16/18 infections for all vaccines regardless of age of vaccine receipt.^19–21^ Additionally, we assumed the 2-valent vaccine provided cross-protective efficacy of 93.8%, 79.1%, and 82.6% for HPV types 31, 33, and 45, respectively.^22^ The duration of protection for type-specific vaccine efficacy was assumed to be lifelong. We chose to take an optimistic approach by assuming that vaccine efficacy has an age-independent effect on reducing the probability of acquiring a newly acquired or reactivated latent infection, but has no effect on prevalent infections or clearance of infections on the causal pathway to cancer. Therefore, as the cumulative proportion of causal infections increases with age, the effectiveness of the vaccine against cancer remains age-dependent. The impact of age-specific vaccine efficacy against an incident infection was explored in sensitivity analysis. Vaccination was applied irrespective of current HPV status, i.e., vaccination was not paired with a primary HPV screening for this exploratory analysis.

**Figure 1.**
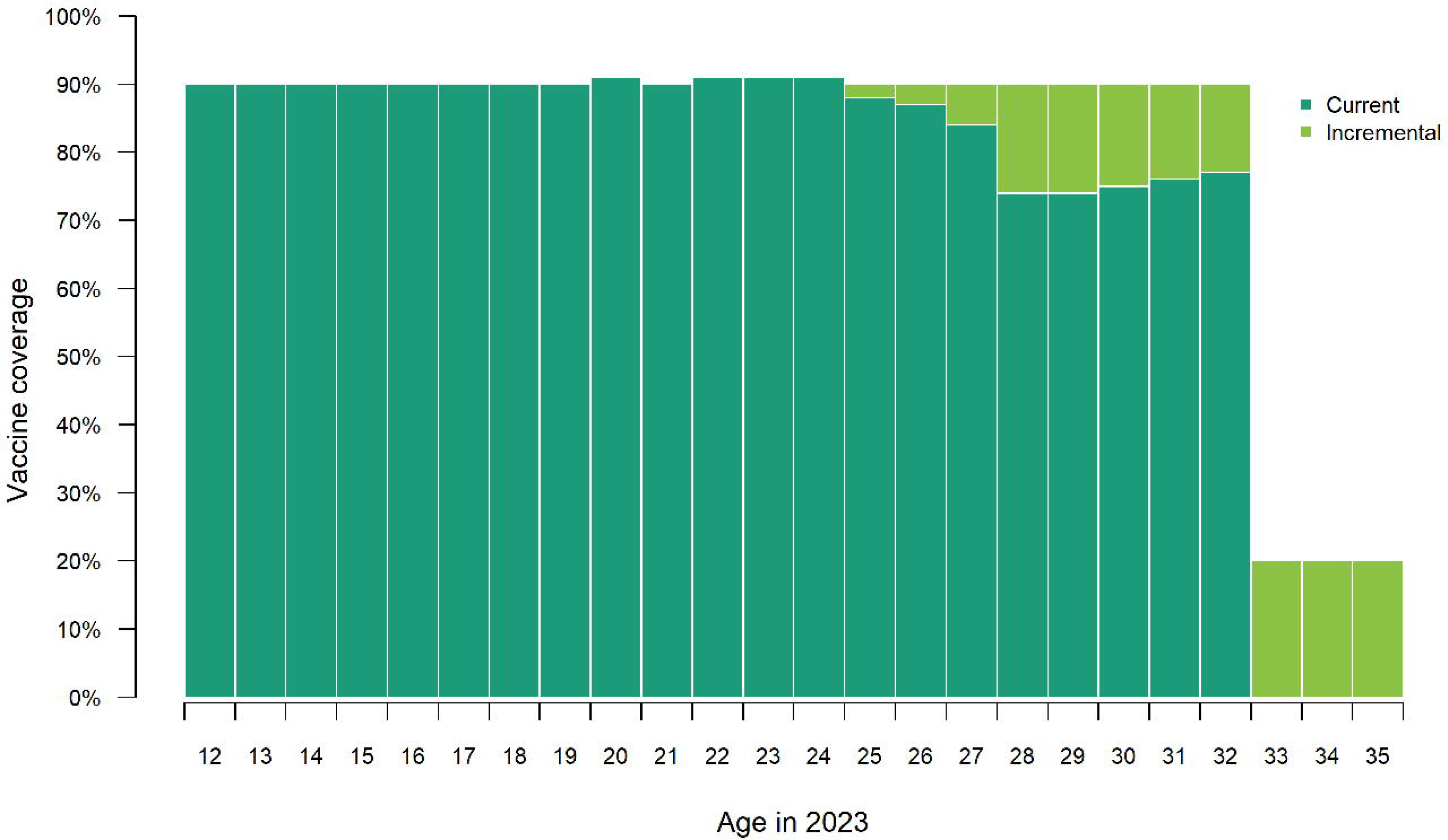
Current levels of routine vaccination coverage for female cohorts aged 12 to 24 and incremental vaccination coverage for female cohorts aged 25 to 35 and in 2023.

To explore the impact of vaccination and de-intensified screening, each vaccination scenario was coupled with a screening scenario that varied the frequency of HPV-based screening (5-yearly (status-quo), 7-yearly, or 10-yearly) for a total of 34 strategies including status-quo. To reflect the newly implemented 2023 screening guidelines in Norway that uniformly recommends HPV-based screening, i.e., replaces 3-yearly cytology with 5-yearly HPV testing for women aged 25– 33 years,^23^ the status-quo screening program assumed 5-yearly primary HPV-based screening for women aged 25 years beginning in 2023. For women aged 26–33 years, we assumed their next eligible primary screening involved primary HPV testing. For all women aged 34–69 years in 2020, we assumed they were screened using primary HPV testing (reflecting the replacement of cytology screening with primary HPV screening in Norway for older women). The full list of analyzed scenarios is detailed in Table 1.

**Table 1.** Analyzed strategies.

| Scenario | Vaccination coverage in 2023 | Screening frequency |
| --- | --- | --- |
| Status-Quo | Status-Quo | 5y |
| Age25 | Incremental coverage of ages 12–25 to max coverage* | 5y; 7y; 10y |
| Age26 | Incremental coverage of ages 12–26 to max coverage* | 5y; 7y; 10y |
| Age27 | Incremental coverage of ages 12–27 to max coverage* | 5y; 7y; 10y |
| Age28 | Incremental coverage of ages 12–28 to max coverage* | 5y; 7y; 10y |
| Age29 | Incremental coverage of ages 12–29 to max coverage* | 5y; 7y; 10y |
| Age30 | Incremental coverage of ages 12–30 to max coverage* | 5y; 7y; 10y |
| Age31 | Incremental coverage of ages 12–31 to max coverage* | 5y; 7y; 10y |
| Age32 | Incremental coverage of ages 12–32 to max coverage* | 5y; 7y; 10y |
| Age33 | Incremental coverage of ages 12–32 to max coverage*<br>and incremental coverage of age 33 to 20% ** | 5y; 7y; 10y |
| Age34 | Incremental coverage of ages 12–32 to max coverage*<br>and incremental coverage of ages 33–34 to 20% ** | 5y; 7y; 10y |
| Age35 | Incremental coverage of ages 12–32 to max coverage*<br>and incremental coverage of ages 33–35 to 20% ** | 5y; 7y; 10y |

\*Maximum coverage reached in routine program = 90% for girls, 89% for boys based on 2021 coverage levels.
\*\*Scenarios of 50% coverage and maximum coverage reached for vaccine-naïve cohorts examined in sensitivity analysis.
Note: status-quo = 5-yearly primary human papillomavirus screening for women aged 25–69 years and vaccination coverage based on 2021 coverage levels; y = yearly.

All scenarios were conducted in the context of imperfect screening coverage centered around the 5-yearly screening interval based on empirical data from the Norwegian screening program.^5,11,24,25^ We assumed that Norwegian women would over-screen at a proportion of 28.2% (3-yearly), under-screen at a proportion of 15% (10–15-yearly), or never attend screening at a proportion of 6%, with the proportion of screening-compliant women (50.8%) centered around the 5-yearly interval. For both the 7-yearly and 10-yearly intervals, we assumed an over-screen proportion of 28.2% (5-yearly), under-screen proportion of 11.3% (15-yearly), never-screen proportion of 6%, and screening-compliant proportion of 54.5% (7-yearly and 10-yearly, respectively).

### Analysis

In order to unmask potentially important cohort-specific differences, we enumerated both population- and cohort-level outcomes. For population-level outcomes, we estimated the health impact on cervical cancer burden in terms of the following outcomes: ASRs of cervical cancer incidence per 100,000 woman-years between 2009 and 2110 (inclusive) and age-standardized HPV-16 prevalence.^26^ For population-level outcomes, model outcomes were aggregated over multiple birth cohorts to capture the lifetime health benefits of women born prior to the year 2110. We additionally estimated the number of lifetime colposcopies performed as a non-health effect and resources use. Importantly, the level of status-quo vaccination coverage varied greatly between the cohorts, e.g., status-quo for the 25-year-old women in 2023 included a very high baseline level of vaccination coverage of 88%, while the status-quo for the 35-year-old women in 2023 did not include any directly protected women. Therefore, we also compared the cohort-specific outcomes to a screening-only, no-vaccination scenario as a standardized comparator to benchmark health benefits under deintensified screening.

We defined the elimination year as the year in which ASR of cervical cancer incidence consistently decreased to <4 new cases per 100,000 woman-years. Base-case results were age-standardized to the standard Norway population (0–84 years).^27^ We calculated the number of colposcopies performed each year by applying the Norway female population projections for 2009 to 2110 from United Nations World Population Prospects.^28^ We extracted the lifetime risk of cervical cancer for cohorts aged 25, 30, and 35 years in 2023 and calculated percentage change in lifetime risk compared to status-quo and a screening-only, no-vaccination scenario.

As the HPV ‘EVEN FASTER’ concept combines the goal of accelerating elimination^2^ with the ‘HPV-FASTER’ concept of reducing the need for frequent screening,^1^ we combined the metrics of elimination year and ASR by 2050 as a proxy metric for monitoring the health impact of reducing screening intensity to evaluate our analytic scenarios. Specifically, we compared to a projected cervical cancer elimination year of 2038 and ASR of 2.29 cervical cancer cases per 100,000 women by 2050 in the status-quo scenario. We categorized the 34 scenarios into one of five groups depending on potential outcomes: (1) scenarios that did not de-intensify screening, decreased ASR in year 2050, but did not accelerate cervical cancer elimination compared to status-quo; (2) strategies that did not de-intensify screening, decreased ASR in year 2050, and accelerated cervical cancer elimination compared to status-quo; (3) strategies that de-intensified screening, decreased ASR in year 2050, but did not accelerate cervical cancer elimination compared to status-quo; (4) strategies that de-intensified screening, decreased ASR in year 2050, and accelerated cervical cancer elimination compared to status-quo; and (5) strategies that de-intensified screening and neither decreased ASR in year 2050 nor accelerated cervical cancer elimination compared to status-quo. Scenarios falling in group 4 would be considered confirmations of the HPV ‘EVEN FASTER’ concept.

### Alternative to ‘EVEN FASTER’ concept

As prior analyses have shown that scaling up screening participation can accelerate cervical cancer elimination,^6^ we also examined an alternative to the ‘EVEN FASTER’ concept which assumed increased screening adherence rather than increased vaccination coverage. We defined this alternative concept to screening adherence in two ways: (1) all women who currently under-screen (15% of women at a 10–15-yearly interval) are adherent with a 5-yearly screening interval; and (2) all women who under-screen and 50% of women who never screen (6% of women) are adherent with a 5-yearly interval. Screening adherence changes were operationalized with two different approaches: (a) restricted to ages 25–35 only; and (b) implementation for all women aged 25–69. Due to computational restrictions, we assumed that increasing adherence occurred in 2020 for women aged 35–69 and in 2023 for women 25–35 years, alongside the switch to primary HPV-based screening for the relevant cohorts.

### Sensitivity analysis

We conducted sensitivity analyses to assess the effect of certain assumptions on our results. First, we assumed two higher alternatives to vaccine coverage level for vaccine-naïve cohorts (i.e., female cohorts aged 33–35 and male cohorts aged 18–35 in 2023), 50% coverage, and 90% coverage, compared to these cohorts reaching 20% coverage in the base-case. Second, we assessed the effect of the World Female Population 2015 (0–99 years)^29^ as an alternative population structure on the elimination year and for comparison to elimination analyses in other countries as the benchmark population structure in use for global predictions by WHO).^30,31^ Third, to account for the potential of reduced vaccine efficacy by age (also a proxy for reduced efficacy in latent HPV infections), we assumed that vaccine efficacy was reduced by 25% for individuals aged 24 and older, consistent with vaccine-efficacy studies,^32,33^ for both historical vaccination and any ‘EVEN FASTER’ vaccination strategies.

## Results

### Population health impact and elimination timing

Increasing vaccination by vaccinating women up to age 25–35 years is expected to decrease circulation of HPV-16, which confirms the ‘EVEN FASTER’ hypothesis; however, the reductions in prevalence decreased at a decreasing rate as vaccination age increased (Figure 2A). In addition, HPV-16 remains in circulation in the population beyond 2050. However, when isolating HPV-16 prevalence only among cohorts aged 25–35, prevalence is expected to plateau at its lowest level (0.1%) approximately one year earlier than status-quo, i.e., without ‘EVEN FASTER’ campaigns (Figure 2B).

**Figure 2.**
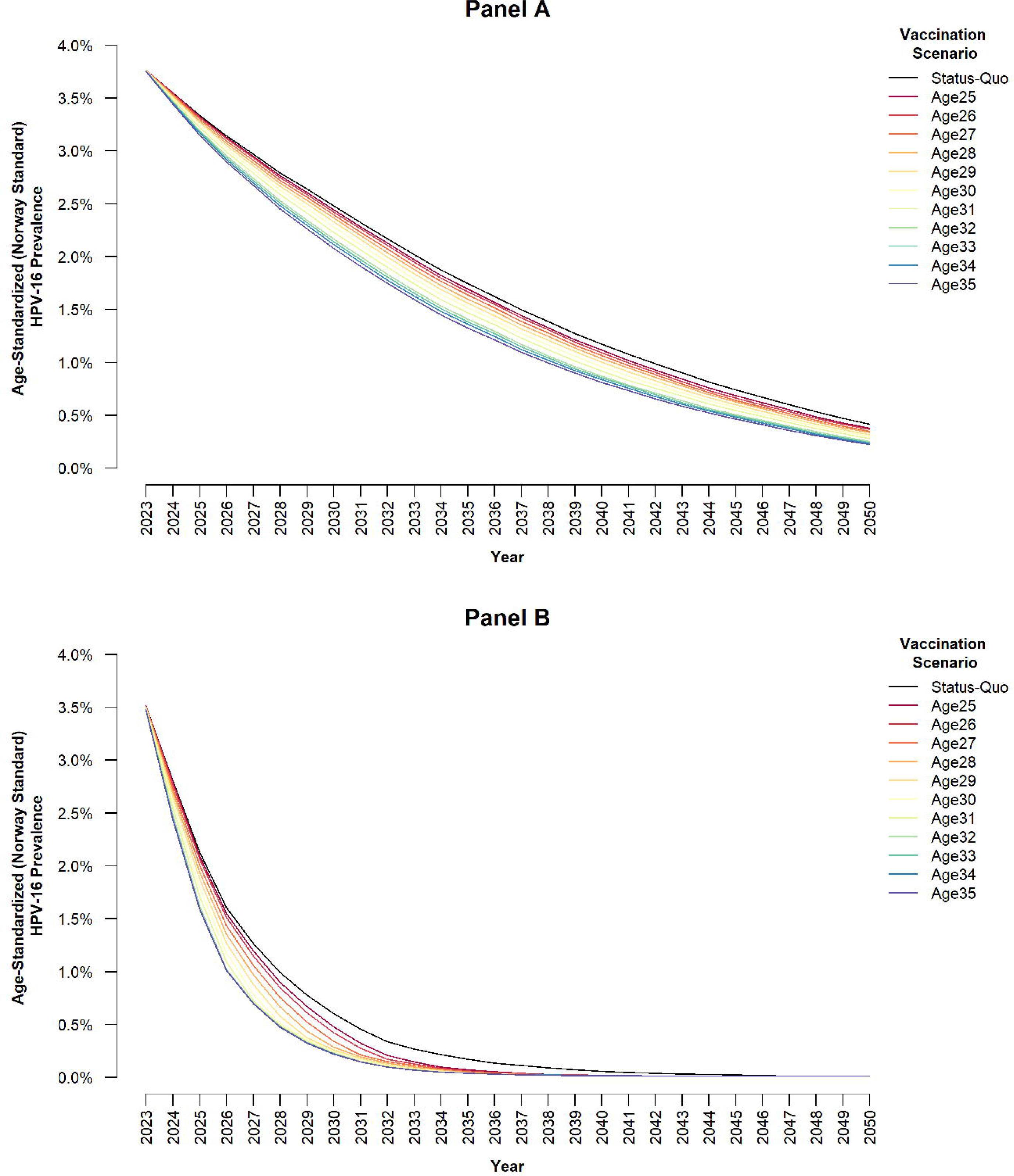
Age-standardized human papillomavirus (HPV) type 16 prevalence associated with ‘EVEN FASTER’ vaccination-only scenarios in Norway for women aged 25–69 years in 2023 (Panel A) and women aged 25–35 years in 2023 (Panel B) Note: Each line represents the greatest cohort age incrementally vaccinated in an ‘EVEN FASTER’ campaign in 2023 (e.g., the “Age26” scenario involved incrementally vaccinating both the age-25 cohort and the age-26 cohort).

With the recent introduction of universal primary HPV testing for all screening-age women in 2023, Norway is projected to accelerate their elimination timeframe by one year (from 2039 to 2038) compared with previous screening guidelines of 3-yearly primary cytology-based screening for women aged 25–33 years followed by a switch to 5-yearly primary HPV testing for women aged 34–69 years (Figure S2). The 2023 guideline also reached a lower cervical cancer incidence of 2.29/100,000 woman-years as compared with the previous prevention policies (2.51/100,000 woman-years).

Under ‘EVEN FASTER’ vaccination campaigns for men and women aged ≤35 years, increasing vaccination coverage by ≤20% coverage (depending on a cohorts’ prior coverage) accelerated the elimination timeframe by one additional year, to 2037, compared to the status-quo elimination year of 2038 (Figure 3A). However, the incremental vaccine coverage would need to be increased for both men and women up to at least age 34 years in order to achieve acceleration of cervical cancer elimination by 1 year (Table S1). All strategies that increased vaccination only (i.e., screening maintained a 5-yearly frequency) resulted in an ASR that was lower than status-quo. When we coupled the vaccination strategies with de-intensified screening, we found 13 strategies—that generally included primary HPV testing at 7-yearly or 10-yearly intervals alongside vaccination of a greater number of cohorts—lowered ASR by 2050 (Figure 3B). We identified a total of 22 strategies that yielded an ASR that was lower than status-quo but did not accelerate cervical cancer elimination (Groups 1 and 3), while no strategies both accelerated cervical cancer elimination and de-intensified screening (Group 4). In other words, our base-case scenario did not support the ‘EVEN FASTER’ concept in terms of projected acceleration of the elimination timeline.

**Figure 3.**
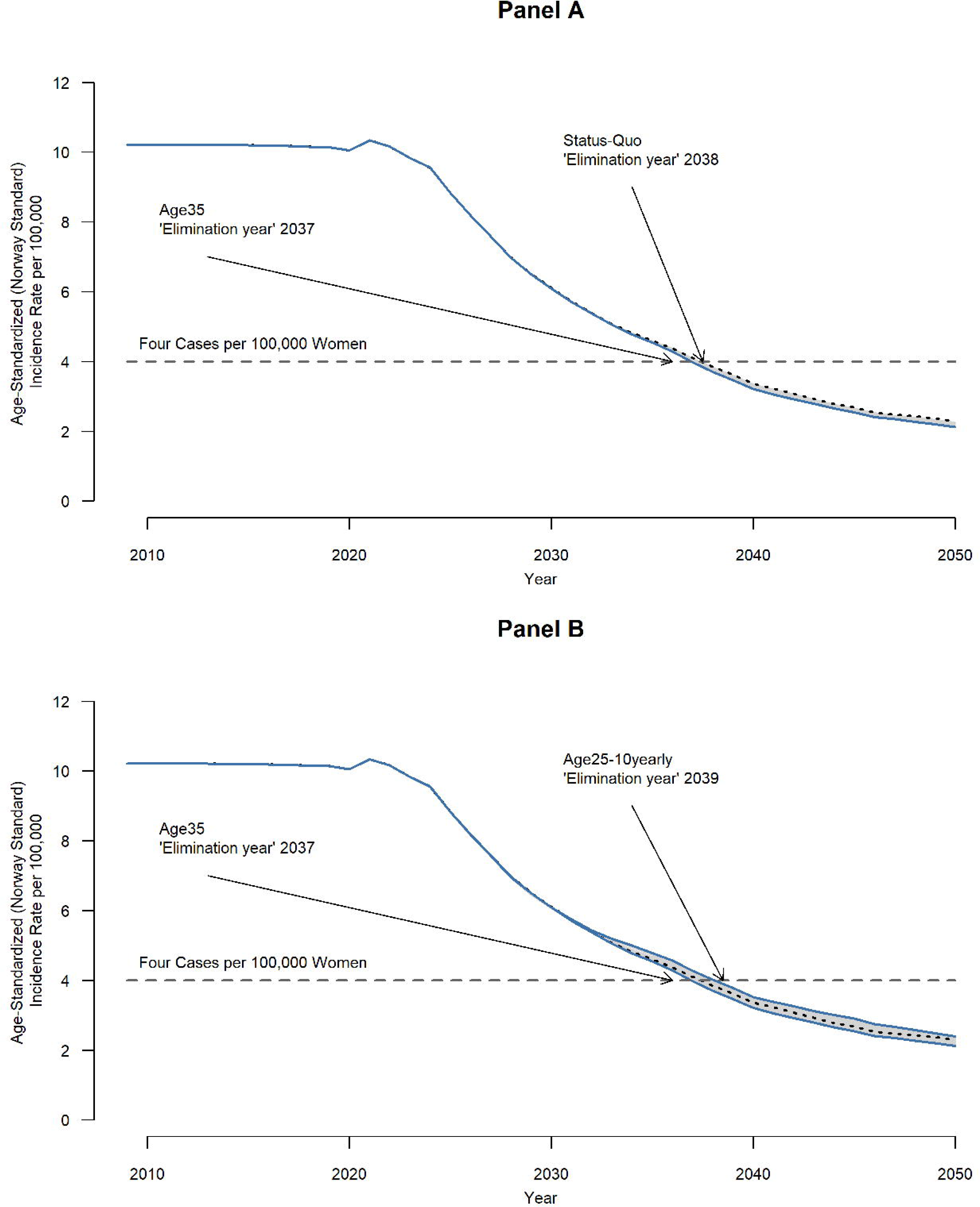
Time to cervical cancer elimination associated with ‘EVEN FASTER’ vaccination scenarios (Panel A) and vaccination + screening scenarios (Panel B) in Norway. Note: The status-quo scenario was estimated to eliminate cervical cancer as a public health problem in 2038.

### Alternative to ‘EVEN FASTER’ concept

When we explored the impact of increasing screening adherence as an alternative to increasing vaccination coverage for women aged 25–35 years, we found elimination timing could be accelerated by one year, but only if both under-screeners and prior never-screeners were reached. A targeted approached to improving adherence among all screen-eligible women (aged 25–69 years) was projected to accelerate elimination between 2 and 5 years depending on the program’s ability to reach never-screeners. Importantly, all screening-adherence related scenarios improved ASR compared to status-quo.

### Cohort-specific health impact

For cohorts aged 25, 30, and 35 years in 2023, all examined strategies resulted in a decreased number of colposcopies compared to status-quo; however, a larger number of colposcopies were avoided as screening frequency decreased and as vaccination age increased (Table 2). In contrast to resource use, the direction of the health effects varied with the strategy. For example, for women aged 25 years in 2023, strategies that incrementally increased vaccination coverage only (i.e., maintained 5-yearly screening frequency) decreased the lifetime risk of cervical cancer by 3.7% compared with cohort-specific status-quo, but screening intensity could only be reduced from 5-yearly to 7-yearly without increasing lifetime risk for this cohort. Although the lifetime risk of cervical cancer for this cohort decreased by 8.7% as vaccination age increased to age 35 years (due to herd immunity), decreasing screening frequency to 10-yearly consistently increased lifetime risk compared to status-quo. However, when changing the comparator, screening could be de-intensified to 10-yearly for 25-year-olds while achieving nearly a 66% reduction in lifetime risk of developing cervical cancer compared with a screening-only (no-vaccination) scenario.

**Table 2.** Lifetime outcomes for women by selected cohort age (25, 30, 35) in 2023*.

| Scenario | Colposcopies avoided<br>compared to status-quo |  |  | Change in lifetime cancer risk<br>compared to status-quo** |  |  | Change in lifetime cancer risk<br>compared to a screening-only,<br>no-vaccination scenario** |  |  |
| --- | --- | --- | --- | --- | --- | --- | --- | --- | --- |
|  | 25 | 30 | 35 | 25 | 30 | 35 | 25 | 30 | 35 |
| Vax up to age 25, 10-yearly screening | 2,824 | 2,385 | 3,158 | 9.1% | 5.3% | 6.3% | -65.5% | -29.6% | -7.4% |
| Vax up to age 25, 7-yearly screening | 1,918 | 1,447 | 2,118 | 0.9% | 1.4% | 2.4% | -68.1% | -32.1% | -10.9% |
| Vax up to age 25, 5-yearly screening | 441 | 163 | 73 | -3.7% | -0.6% | -0.1% | -69.6% | -33.5% | -13.0% |
| Vax up to age 30, 10-yearly screening | 3,198 | 3,800 | 3,594 | 3.8% | -2.9% | 4.6% | -67.2% | -35.1% | -8.9% |
| Vax up to age 30, 7-yearly screening | 2,354 | 3,096 | 2,623 | -3.3% | -6.2% | 0.8% | -69.5% | -37.3% | -12.2% |
| Vax up to age 30, 5-yearly screening | 978 | 2,169 | 702 | -8.1% | -7.2% | -1.4% | -71.0% | -37.9% | -14.2% |
| Vax up to age 35, 10-yearly screening | 3,296 | 3,954 | 4,943 | 2.6% | -3.7% | -1.7% | -67.6% | -35.6% | -14.4% |
| Vax up to age 35, 7-yearly screening | 2,466 | 3,270 | 4,173 | -4.2% | -6.8% | -4.9% | -69.7% | -37.7% | -17.2% |
| Vax up to age 35, 5-yearly screening | 1,111 | 2,385 | 2,636 | -8.7% | -7.8% | -7% | -71.2% | -38.4% | -19.0% |

\* Scaled to estimated number of women in Norway for each cohort age in 2023: 35,702 women aged 25; 37,160 women aged 30; 37,800 women aged 35.
\*\* Positive value indicates increase in lifetime cancer risk and negative value indicates decrease in lifetime cancer risk vs comparator scenario.
Note: Vax = vaccinate incrementally to maximum achieved vaccination coverage for previously vaccinated cohorts (females aged 12–32 and males aged 12–17 in 2023) and to 20% coverage for vaccine-naïve cohorts (females aged 33–35 and males aged 18–35 in 2023). Status-quo lifetime cervical cancer risk was 0.175%, 0.437%, and 0.588% for cohorts aged 25, 30, and 35 in 2023, respectively. Screening-only, no-vaccination scenario lifetime cervical cancer risk was 0.555%, 0.653%, and 0.675% for cohorts aged 25, 30, and 35 in 2023, respectively.

For women aged 30 years in 2023, increasing vaccination coverage up to age 30 years yields the greatest health impact by maintaining the 5-yearly screening frequency (7.2% reduction in lifetime risk compared to status-quo), but this cohort can reduce screening frequency to 10-yearly and continue to maintain health benefits (2.9% reduction in lifetime risk compared to status-quo). Similarly for women aged 35 in 2023, increasing vaccination coverage up to age 35 still allows for concomitantly decreasing screening frequency with improved lifetime health outcomes compared to status-quo. The projected reductions in lifetime risk were greater when compared to a screening-only, no-vaccination (and no circulating herd immunity) scenario.

### Sensitivity analysis

When we assumed that vaccine-naïve cohorts (i.e., female cohorts aged 33–35 years and male cohorts aged 18–35 years in 2023) could achieve 50% coverage (rather than 20%), we still did not identify any strategies that both accelerated cervical cancer elimination and de-intensified screening (Table S2). However, when we assumed that vaccine-naïve cohorts reached maximum-achieved coverage (i.e., 90% for females and 89% for males), we identified in two strategies that were projected to both accelerate cervical cancer elimination and de-intensify screening: vaccinating up to age 34 or age 35 with a 7-yearly screening frequency (Table S3). For example, compared to 20% coverage in the base-case scenario, assuming that vaccine-naïve cohorts achieved 50% coverage resulted in greater reductions in the lifetime risk of cervical cancer compared to status-quo: 11.4% for 5-yearly screening, 9.6% for 7-yearly screening, and 6.7% for 10-yearly screening for the age 35 cohort in 2023 (Table S4). Similarly, assuming that vaccine-naïve cohorts achieved maximum-achieved coverage maximized the reductions in the lifetime risk of cervical cancer compared to status-quo: 15.3% for 5-yearly screening, 13.9% for 7-yearly screening, and 11.2% for 10-yearly screening for the age 35 cohort in 2023 (Table S4).

Compared to the optimistic full vaccine efficacy in the base-case scenario, assuming that vaccine efficacy was reduced by 25% for women aged 24 and older, for both historical vaccination and any additional ‘EVEN FASTER’ vaccination, resulted in a projected elimination year of 2038 or later for all examined scenarios (i.e., no strategies in Groups 2 and 4) (Table S5). Among the 18 strategies that decreased ASR, 7 strategies also involved a de-intensified screening frequency of 7-yearly screening but required vaccination up to at least age 29 years. From an individual cohort perspective, the overall benefits of vaccination decreased and there was more uncertainty in the potential to simultaneously reduce the screening frequency (Table S6). For example, increasing vaccination to age 35 years and reducing screening frequency to 10-yearly no longer provided reductions in lifetime cancer risk compared with status quo for the 35-year-olds. Finally, compared to a projected elimination year of 2038 given age-standardization with the Norway standard population in the base-case scenario, age-standardization with the World Female Population 2015 resulted in a projected elimination year of 2034 or later for all examined scenarios (i.e., no strategies in Groups 2 and 4) (Table S7).

## Discussion

In our analysis, we tested the hypotheses that HPV ‘EVEN FASTER’ could accelerate cervical cancer elimination and reduce the need for screening. We also compared the ‘EVEN FASTER’ vaccination-based interventions with screening-based interventions to achieve the same endpoints. Although we projected that 22 different analyzed strategies would yield an ASR that was lower than status-quo from a population perspective, none of the analyzed strategies both de-intensified screening and accelerated the timeline for cervical cancer to be eliminated as a public health problem, which does not support the proposed benefits of the ‘EVEN FASTER’ concept. Two strategies did accelerate the timeline for elimination, involving incremental vaccine coverage added for all women up to age 34 or 35 with current screening intensity (5-yearly) maintained, resulting in a projected elimination year of 2037 (compared to 2038 in status-quo). However, vaccinating cohorts up to age 35 (assuming ≤20% coverage for vaccine-naïve cohorts) would involve administering 377,149 additional doses of HPV vaccine (to both males and females, assuming a 3-dose schedule for individuals aged 15 years and older) and cost more than $141 billion USD (assuming a cost of $375 USD per person). In sensitivity analysis, we identified at least one strategy that confirmed the ‘EVEN FASTER’ hypothesis, but would require vaccination of vaccine-naïve cohorts at maximum-achieved coverage levels (i.e., 90% for females and 89% for males).

As an alternative to vaccine-based interventions to increase cervical cancer elimination according to the ‘EVEN FASTER’ concept, we looked at scenarios that increased the levels of adherence to screening and projected elimination timeframes that could be as early as the year 2033. Importantly, we projected that increases to screening adherence achieved the greatest health impact in the long term (ASR by 2050 of 1.62–2.10/100,000 woman-years for the most optimistically increased screening adherence compared to 2.12/100,000 woman-years for the most optimistically increased vaccination coverage). Compared to clinician-collected HPV testing, self-collected HPV testing approaches have been shown to be effective and cost-effective.^10,34,35^ A previous analysis of cervical cancer elimination in the United States has also shown that scaling up screening participation is a viable option to accelerate the timeline to elimination^6^ and is likely considered cost-effective in Norway.^36,37^ To move the needle on the elimination timeframe, comprehensive strategies that target women at ages where the burden of cervical cancer is greatest (ages 35–50 years in 2023), are required. Whether incremental vaccination coverage or increased screening adherence influence projected elimination timeframes, all examined scenarios in this analysis are time-sensitive as cohorts continue to age (and experience their causal HPV infection).

There were some important differences in our results between our population-level outcomes and our cohort-specific outcomes. For example, at a population level, we found that vaccinating at least to age 29 or 30 years enabled reductions in screening frequency (Group 3); however, at a cohort level, some reduced screening frequencies implied increases in lifetime risk for the age 25 cohort (when compared with status quo). In addition, we found seemingly counterintuitive results that screening frequency could not be reduced to 10-yearly screening and reduce the lifetime risk of cancer among the 25-year-olds but could be reduced to 10-yearly screening for the two older cohorts. As nearly 88% of women aged 25 in 2023 were vaccinated prior to the ‘EVEN FASTER’ campaign scenarios analyzed, the ‘EVEN FASTER’ campaign implied only an approximately 2% increase in coverage for women, which yields a relatively small marginal increase in the status-quo benefits. Consequently, and despite these women having the lowest status-quo lifetime cervical cancer risk (i.e., 0.175%), decreasing screening frequency under small incremental increases in coverage led to increases in lifetime risk when compared with their own status-quo benefits. Additionally, this cohort only became eligible for screening in 2023, whereas older cohorts that previously entered the screening program may have received protection from prior lifetime screens at 5-yearly screening intervals compared to the 25-year-old cohort. As previously mentioned, the benchmark against which we measure health improvements can be a moving target for vaccinated or partially-vaccinated cohorts. When compared against a more standardized comparator, i.e., a screening-only and no-vaccination scenario, the 25-year-olds can be screened less frequently while experiencing a nearly 66% reduction in lifetime risk, which may be important to maintain the harm-benefits ratios of screening.^11^

We conducted an exploratory analysis looking at both feasible and maximum-achievable benefits of an ‘EVEN FASTER’ vaccination policy, but we did not assume that vaccination was paired with an initial primary HPV screen. In other words, we applied vaccination irrespective of HPV status as proposed in a recent white paper by the Cancer Registry of Norway.^15^ However, compared to the white paper’s proposal, we neither explored the use of 9-valent vaccine, nor re-vaccination of women previously vaccinated with the 2-valent vaccine. As the majority of cervical cancers that occur among younger women (the target of ‘EVEN FASTER’ campaigns) are caused by HPV-16 and HPV-18,^38^ of which both types are already directly protected by the 2-valent vaccine, we do not expect the 9-valent to affect the timeframe to elimination substantially. In a prior analysis,^5^ we found that the 9-valent vaccine yielded an incremental benefit compared with the 2-valent; however, these incremental benefits were not considered cost-effective in the routine childhood vaccination program unless there were large discounts on the vaccine price. The prior cost-effectiveness analysis also included the added benefits the 9-valent vaccine types may have on preventing screening-positives and precancer treatments, but these outcomes were not considered in this health impact analysis.

There are several limitations to consider. Given the varied age-specific trends in vaccination coverage, there is much uncertainty in potential achievable coverage for a 1-year vaccination campaign. Therefore, we explored a range of coverage assumptions but assumed a conservative estimate of 20% in the base case, and 50% and 90% in scenario analyses. We made a simplifying assumption that the initial switch to primary HPV screening for women aged 34–69 occurred nationwide in 2020 rather than gradual scale-up over multiple years and that the switch to primary HPV screening occurred immediately for women aged 25 and older in 2023 who had not previously switched. The exact timing of elimination of the new ‘status quo’ screening program in Norway is likely impacted by these assumptions. We also did not consider explicit reactivation of latent infections and potential differences in vaccine efficacy for reactivated infections, potentially overestimating vaccine benefit, but we included a scenario that assumed a 25% reduction in vaccine efficacy for all individuals aged 24 years and older as a proxy. We did not consider the impact of ‘EVEN FASTER’ vaccination campaigns on non-cervical cancers; however, there are similar considerations around the proportion of men and women already infected with the causal HPV infections that will lead a non-cervical cancer and potential age-specific effects on efficacy for these individuals. Finally, we did not consider the potential correlation between women who are considered ‘over-screeners’ according to current Norwegian screening guidelines and who may then opt in for ‘EVEN FASTER’ vaccination, which may reduce the overall impact of a vaccination campaign.

Finally, whether or not the ‘EVEN FASTER’ approach would be considered cost-effective according to priority-setting guidelines in Norway needs to be evaluated. In Norway, priority-setting is determined according to both health impact, such as ASR and/or the timing of cervical cancer elimination and the severity of the disease, and the economic efficiency, such as with a cost-effectiveness analysis framework. In addition, as the proposed ‘EVEN FASTER’ concept requires a large allocation of global vaccine supply resources, global vaccine equity considerations might need to be considered when prioritizing interventions to accelerate cervical cancer elimination in Norway.

Increasing vaccination paired with less intensive screening frequencies can lead to greater benefits but would not accelerate cervical cancer elimination compared to current prevention policies. In contrast, interventions targeting under- and never-screeners may both eliminate cervical cancer faster and lead to greater cervical cancer health benefits compared with vaccination-based interventions. Future evaluations of HPV ‘EVEN FASTER’ will need to estimate the full economic implications of these policies. The next step to determine the best combination of strategies would be to conduct a full cost-effectiveness analysis to see if the health gains would be worth the costs.

## Additional Information

## Acknowledgements

The authors thank Bo T. Hansen, Berit Feiring, Ida Laake, Mari Nygård, Lill Trogstad, Megan A. Smith, and Stephen Sy for their collaboration and support on the previous analysis that this work builds on.

## Authors’ contributions

AP and EAB conceptualized the study, conducted the data analysis, and drafted the initial manuscript. KP and JJK critically reviewed the analysis and manuscript. All authors approved the final version of the manuscript.

## Ethics approval and consent to participate

Not applicable.

## Consent for publication

Not applicable.

## Data availability

The datasets generated during and/or analyzed during the current study are available from the corresponding author on reasonable request.

