## Supplementary Appendix for "Impact of ‘EVEN FASTER’ concept to accelerate cervical cancer elimination in Norway: A model-based analysis"

### Figure S1. Current levels of routine vaccination coverage for male cohorts aged 12 to 17 and incremental vaccination coverage for male cohorts aged 18 to 35 and in 2023

Vaccination coverage

| Age in 2023 | Current | Incremental |
| --- | --- | --- |
| 12 | 89% | 0% |
| 13 | 89% | 0% |
| 14 | 89% | 0% |
| 15 | 89% | 0% |
| 16 | 89% | 0% |
| 17 | 89% | 0% |
| 18 | 0% | 20% |
| 19 | 0% | 20% |
| 20 | 0% | 20% |
| 21 | 0% | 20% |
| 22 | 0% | 20% |
| 23 | 0% | 20% |
| 24 | 0% | 20% |
| 25 | 0% | 20% |
| 26 | 0% | 20% |
| 27 | 0% | 20% |
| 28 | 0% | 20% |
| 29 | 0% | 20% |
| 30 | 0% | 20% |
| 31 | 0% | 20% |
| 32 | 0% | 20% |
| 33 | 0% | 20% |
| 34 | 0% | 20% |
| 35 | 0% | 20% |

### Figure S2. Impact of 2023 guidelines on time to cervical cancer elimination in Norway

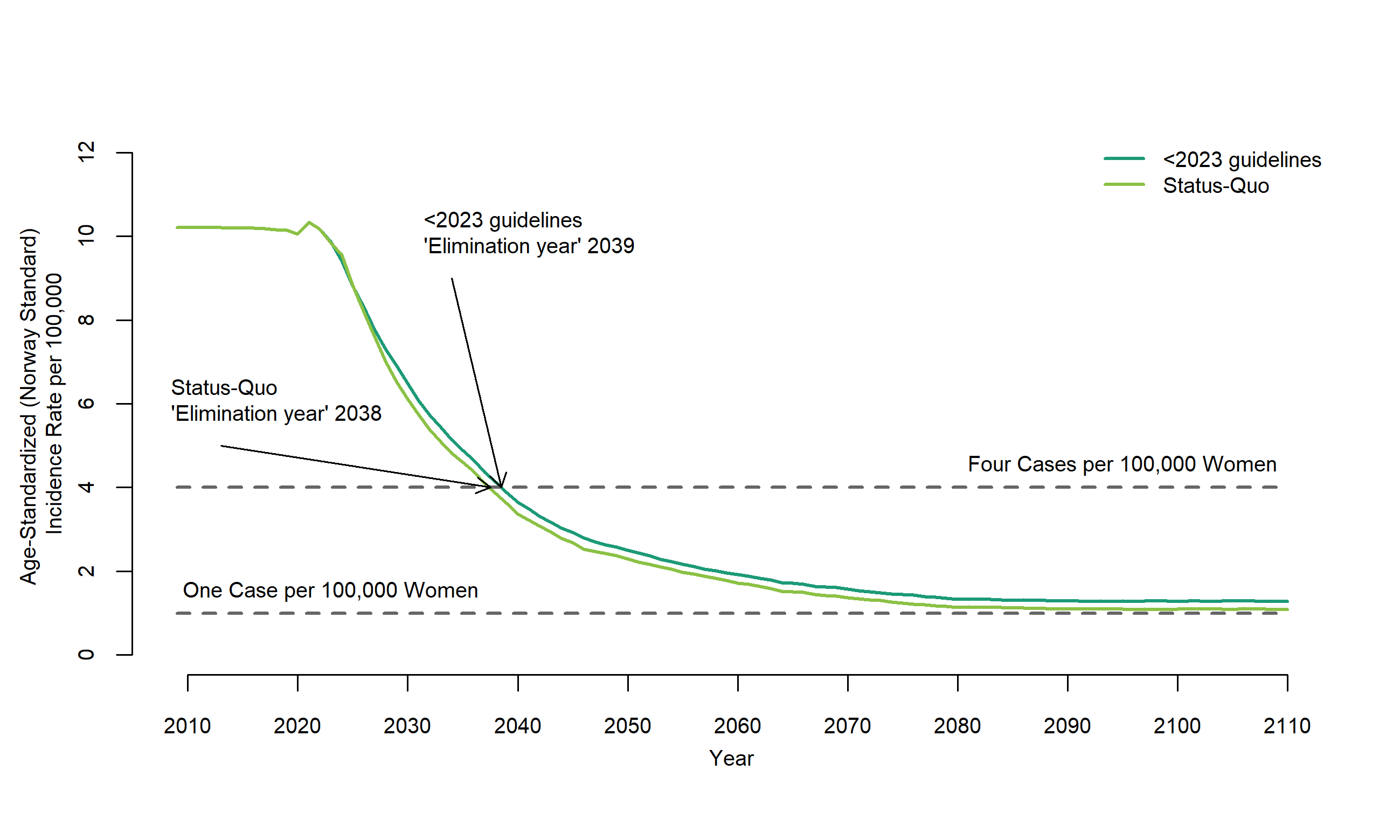

Note: <2023 guidelines = Switched from cytology to primary HPV-based testing at age 34 years in 2020; Status-Quo (≥July 2023 guidelines) = Switched to primary HPV-based testing beginning at age 25 in 2023, status-quo in Norway.

### Table S1. Elimination timeframe and age-standardized incidence rate associated with ‘EVEN FASTER’ scenarios in Norway, assuming 20% coverage for vaccine-naïve cohorts (females aged 33–35 and males aged 18–35)

| Vaccinate at maximum coverage up to specified age in 2023 | Screening frequency (years) | Year elimination (<4 cases per 100,000 women) achieved | Age-standardized incidence rate (cases per 100,000 women) in year 2050 |
| --- | --- | --- | --- |
| Status-Quo | 5 | 2038 | 2.290 |
| **Group 1. Vaccination-only strategies that decreased age-standardized incidence rate in year 2050 but did not accelerate cervical cancer elimination compared to status-quo** | | | |
| 25 | 5 | 2038 | 2.262 |
| 26 | 5 | 2038 | 2.252 |
| 27 | 5 | 2038 | 2.242 |
| 28 | 5 | 2038 | 2.219 |
| 29 | 5 | 2038 | 2.206 |
| 30 | 5 | 2038 | 2.190 |
| 31 | 5 | 2038 | 2.180 |
| 32 | 5 | 2038 | 2.148 |
| 33 | 5 | 2038 | 2.141 |
| **Group 2. Vaccination-only strategies that decreased age-standardized incidence rate in year 2050 and accelerated cervical cancer elimination compared to status-quo** | | | |
| 34 | 5 | 2037 | 2.119 |
| 35 | 5 | 2037 | 2.116 |
| **Group 3. Strategies that de-intensified screening and decreased age-standardized incidence rate in year 2050, but did not accelerate cervical cancer elimination compared to status-quo** | | | |
| 31 | 10 | 2038 | 2.287 |
| 32 | 10 | 2038 | 2.287 |
| 33 | 10 | 2038 | 2.280 |
| 28 | 7 | 2038 | 2.271 |
| 34 | 10 | 2038 | 2.271 |
| 35 | 10 | 2038 | 2.256 |
| 29 | 7 | 2038 | 2.253 |
| 30 | 7 | 2038 | 2.233 |
| 31 | 7 | 2038 | 2.227 |
| 32 | 7 | 2038 | 2.197 |
| 33 | 7 | 2038 | 2.196 |
| 34 | 7 | 2038 | 2.179 |
| 35 | 7 | 2038 | 2.160 |
| **Group 4. Strategies that de-intensified screening, decreased age-standardized incidence rate in year 2050, and accelerated cervical cancer elimination compared to status-quo compared to status-quo** | | | |
| **--** | **--** | **--** | **--** |
| **Group 5. Strategies that de-intensified screening, did not decrease age-standardized incidence rate in year 2050, and did not accelerate cervical cancer elimination compared to status-quo** | | | |
| 25 | 10 | 2039 | 2.396 |
| 26 | 10 | 2039 | 2.386 |
| 27 | 10 | 2038 | 2.370 |
| 28 | 10 | 2038 | 2.353 |
| 29 | 10 | 2038 | 2.335 |
| 30 | 10 | 2038 | 2.322 |
| 25 | 7 | 2038 | 2.307 |
| 26 | 7 | 2038 | 2.298 |
| 27 | 7 | 2038 | 2.290 |

Note: Strategies organized in order of decreasing age-standardized incidence rate within group. Heat map formatting for age-standardized incidence rate shows lower cervical cancer incidence rates in green and higher rates in red compared to the rate estimated under existing prevention policies (2.290 cases per 100,000 women in year 2050).

### Table S2. Elimination timeframe and age-standardized incidence rate associated with ‘EVEN FASTER’ scenarios in Norway, assuming 50% coverage for vaccine-naïve cohorts (females aged 33–35 and males aged 18–35)

| Vaccinate at maximum coverage up to specified age in 2023 | Screening frequency (years) | Year elimination (<4 cases per 100,000 women) achieved | Age-standardized incidence rate (cases per 100,000 women) in year 2050 |
| --- | --- | --- | --- |
| Status-Quo | 5 | 2038 | 2.290 |
| **Group 1. Vaccination-only strategies that decreased age-standardized incidence rate in year 2050 but did not accelerate cervical cancer elimination compared to status-quo** | | | |
| 25 | 5 | 2038 | 2.237 |
| 26 | 5 | 2038 | 2.227 |
| 27 | 5 | 2038 | 2.198 |
| 28 | 5 | 2038 | 2.183 |
| 29 | 5 | 2038 | 2.176 |
| 30 | 5 | 2038 | 2.152 |
| 31 | 5 | 2038 | 2.139 |
| 32 | 5 | 2038 | 2.111 |
| **Group 2. Vaccination-only strategies that decreased age-standardized incidence rate in year 2050 and accelerated cervical cancer elimination compared to status-quo** | | | |
| 33 | 5 | 2037 | 2.093 |
| 34 | 5 | 2037 | 2.072 |
| 35 | 5 | 2037 | 2.048 |
| **Group 3. Strategies that de-intensified screening and decreased age-standardized incidence rate in year 2050, but did not accelerate cervical cancer elimination compared to status-quo** | | | |
| 30 | 10 | 2038 | 2.289 |
| 25 | 7 | 2038 | 2.286 |
| 26 | 7 | 2038 | 2.277 |
| 31 | 10 | 2038 | 2.275 |
| 27 | 7 | 2038 | 2.252 |
| 32 | 10 | 2038 | 2.247 |
| 33 | 10 | 2038 | 2.233 |
| 28 | 7 | 2038 | 2.230 |
| 29 | 7 | 2038 | 2.228 |
| 34 | 10 | 2038 | 2.204 |
| 30 | 7 | 2038 | 2.200 |
| 31 | 7 | 2038 | 2.186 |
| 35 | 10 | 2038 | 2.184 |
| 32 | 7 | 2038 | 2.159 |
| 33 | 7 | 2038 | 2.138 |
| 34 | 7 | 2038 | 2.115 |
| 35 | 7 | 2038 | 2.096 |
| **Group 4. Strategies that de-intensified screening, decreased age-standardized incidence rate in year 2050, and accelerated cervical cancer elimination compared to status-quo compared to status-quo** | | | |
| **--** | **--** | **--** | **--** |
| **Group 5. Strategies that de-intensified screening, did not decrease age-standardized incidence rate in year 2050, and did not accelerate cervical cancer elimination compared to status-quo** | | | |
| 25 | 10 | 2038 | 2.364 |
| 26 | 10 | 2038 | 2.363 |
| 27 | 10 | 2038 | 2.330 |
| 28 | 10 | 2038 | 2.317 |
| 29 | 10 | 2038 | 2.312 |

Note: Strategies organized in order of decreasing age-standardized incidence rate within group. Heat map formatting for age-standardized incidence rate shows lower cervical cancer incidence rates in green and higher rates in red compared to the rate estimated under existing prevention policies (2.290 cases per 100,000 women in year 2050).

### Table S3. Elimination timeframe and age-standardized incidence rate associated with ‘EVEN FASTER’ scenarios in Norway, assuming maximum-achieved coverage (90% for females and 89% for males) for vaccine-naïve cohorts (females aged 33–35 and males aged 18–35)

| Vaccinate at maximum coverage up to specified age in 2023 | Screening frequency (years) | Year elimination (<4 cases per 100,000 women) achieved | Age-standardized incidence rate (cases per 100,000 women) in year 2050 |
| --- | --- | --- | --- |
| Status-Quo | 5 | 2038 | 2.290 |
| **Group 1. Vaccination-only strategies that decreased age-standardized incidence rate in year 2050 but did not accelerate cervical cancer elimination compared to status-quo** | | | |
| 25 | 5 | 2038 | 2.207 |
| 26 | 5 | 2038 | 2.192 |
| 27 | 5 | 2038 | 2.173 |
| 28 | 5 | 2038 | 2.154 |
| 29 | 5 | 2038 | 2.140 |
| 30 | 5 | 2038 | 2.118 |
| **Group 2. Vaccination-only strategies that decreased age-standardized incidence rate in year 2050 and accelerated cervical cancer elimination compared to status-quo** | | | |
| 31 | 5 | 2037 | 2.099 |
| 32 | 5 | 2037 | 2.081 |
| 33 | 5 | 2037 | 2.051 |
| 34 | 5 | 2037 | 2.022 |
| 35 | 5 | 2037 | 1.993 |
| **Group 3. Strategies that de-intensified screening and decreased age-standardized incidence rate in year 2050, but did not accelerate cervical cancer elimination compared to status-quo** | | | |
| 28 | 10 | 2038 | 2.281 |
| 29 | 10 | 2038 | 2.276 |
| 25 | 7 | 2038 | 2.254 |
| 26 | 7 | 2038 | 2.252 |
| 30 | 10 | 2038 | 2.250 |
| 27 | 7 | 2038 | 2.234 |
| 31 | 10 | 2038 | 2.230 |
| 32 | 10 | 2038 | 2.213 |
| 28 | 7 | 2038 | 2.201 |
| 29 | 7 | 2038 | 2.189 |
| 33 | 10 | 2038 | 2.180 |
| 30 | 7 | 2038 | 2.170 |
| 31 | 7 | 2038 | 2.143 |
| 34 | 10 | 2038 | 2.143 |
| 32 | 7 | 2038 | 2.127 |
| 35 | 10 | 2038 | 2.112 |
| 33 | 7 | 2038 | 2.094 |
| **Group 4. Strategies that de-intensified screening, decreased age-standardized incidence rate in year 2050, and accelerated cervical cancer elimination compared to status-quo compared to status-quo** | | | |
| 34 | 7 | 2037 | 2.062 |
| 35 | 7 | 2037 | 2.034 |
| **Group 5. Strategies that de-intensified screening, did not decrease age-standardized incidence rate in year 2050, and did not accelerate cervical cancer elimination compared to status-quo** | | | |
| 25 | 10 | 2038 | 2.338 |
| 26 | 10 | 2038 | 2.329 |
| 27 | 10 | 2038 | 2.313 |

Note: Strategies organized in order of decreasing age-standardized incidence rate within group. Heat map formatting for age-standardized incidence rate shows lower cervical cancer incidence rates in green and higher rates in red compared to the rate estimated under existing prevention policies (2.290 cases per 100,000 women in year 2050).

### Table S4. Change in lifetime cancer risk compared to status-quo for women by selected cohort age (25, 30, 35) in 2023

| **Scenario** | **Change in lifetime cancer risk compared to status-quo*** | | | | | | | | |
| --- | --- | --- | --- | --- | --- | --- | --- | --- | --- |
|  | **25** | | | **30** | | | **35** | | |
|  | **20%** | **50%** | **90%** | **20%** | **50%** | **90%** | **20%** | **50%** | **90%** |
| Vax up to age 25, 10-yearly screening | 9.1% | 5.2% | 2.1% | 5.3% | 4.5% | 4.0% | 6.3% | 6.1% | 5.7% |
| Vax up to age 25, 7-yearly screening | 0.9% | -2.3% | -5.3% | 1.4% | 0.8% | 0.5% | 2.4% | 2.3% | 2.0% |
| Vax up to age 25, 5-yearly screening | -3.7% | -6.9% | -9.3% | -0.6% | -1.0% | -1.5% | -0.1% | -0.3% | -0.4% |
| Vax up to age 30, 10-yearly screening | 3.8% | 0.6% | -3.9% | -2.9% | -3.4% | -3.7% | 4.6% | 4.0% | 3.5% |
| Vax up to age 30, 7-yearly screening | -3.3% | -6.5% | -10.5% | -6.2% | -6.7% | -6.9% | 0.8% | 0.1% | -0.3% |
| Vax up to age 30, 5-yearly screening | -8.1% | -10.5% | -14.0% | -7.2% | -7.6% | -7.7% | -1.4% | -1.9% | -2.3% |
| Vax up to age 35, 10-yearly screening | 2.6% | -1.8% | -5.4% | -3.7% | -4.4% | -5.0% | -1.7% | -6.7% | -11.2% |
| Vax up to age 35, 7-yearly screening | -4.2% | -8.2% | -12.0% | -6.8% | -7.5% | -8.3% | -4.9% | -9.6% | -13.9% |
| Vax up to age 35, 5-yearly screening | -8.7% | -12.2% | -15.4% | -7.8% | -8.3% | -8.9% | -7.0% | -11.4% | -15.3% |

* Positive value indicates increase in lifetime cancer risk and negative value indicates decrease in lifetime cancer risk compared to status-quo.

Note: Vax = vaccinate incrementally to maximum achieved vaccination coverage for previously vaccinated cohorts (females aged 12–32 and males aged 12–17 in 2023) and to 20% coverage for vaccine-naïve cohorts (females aged 33–35 and males aged 18–35 in 2023).

### Table S5. Elimination timeframe and age-standardized incidence rate associated with ‘EVEN FASTER’ scenarios in Norway, assuming 25% reduction in vaccine efficacy for cohorts aged 24 years and older

| Vaccinate at maximum coverage up to specified age in 2023 | Screening frequency (years) | Year elimination (<4 cases per 100,000 women) achieved | Age-standardized incidence rate (cases per 100,000 women) in year 2050 |
| --- | --- | --- | --- |
| Status-Quo | 5 | 2038 | 2.330 |
| **Group 1. Vaccination-only strategies that decreased age-standardized incidence rate in year 2050 but did not accelerate cervical cancer elimination compared to status-quo** | | | |
| 25 | 5 | 2038 | 2.317 |
| 26 | 5 | 2038 | 2.297 |
| 27 | 5 | 2038 | 2.296 |
| 28 | 5 | 2038 | 2.285 |
| 29 | 5 | 2038 | 2.267 |
| 30 | 5 | 2038 | 2.256 |
| 31 | 5 | 2038 | 2.245 |
| 32 | 5 | 2038 | 2.225 |
| 33 | 5 | 2038 | 2.212 |
| 34 | 5 | 2038 | 2.199 |
| 35 | 5 | 2038 | 2.195 |
| **Group 2. Vaccination-only strategies that decreased age-standardized incidence rate in year 2050 and accelerated cervical cancer elimination compared to status-quo** | | | |
| **--** | **--** | **--** | **--** |
| **Group 3. Strategies that de-intensified screening and decreased age-standardized incidence rate in year 2050, but did not accelerate cervical cancer elimination compared to status-quo** | | | |
| 29 | 7 | 2038 | 2.324 |
| 30 | 7 | 2038 | 2.310 |
| 31 | 7 | 2038 | 2.291 |
| 32 | 7 | 2038 | 2.272 |
| 33 | 7 | 2038 | 2.260 |
| 34 | 7 | 2038 | 2.246 |
| 35 | 7 | 2038 | 2.252 |
| **Group 4. Strategies that de-intensified screening, decreased age-standardized incidence rate in year 2050, and accelerated cervical cancer elimination compared to status-quo compared to status-quo** | | | |
| **--** | **--** | **--** | **--** |
| **Group 5. Strategies that de-intensified screening, did not decrease age-standardized incidence rate in year 2050, and did not accelerate cervical cancer elimination compared to status-quo** | | | |
| 25 | 10 | 2039 | 2.465 |
| 26 | 10 | 2039 | 2.450 |
| 27 | 10 | 2039 | 2.448 |
| 28 | 10 | 2039 | 2.436 |
| 29 | 10 | 2039 | 2.417 |
| 30 | 10 | 2039 | 2.398 |
| 31 | 10 | 2039 | 2.383 |
| 25 | 7 | 2039 | 2.374 |
| 32 | 10 | 2039 | 2.361 |
| 27 | 7 | 2039 | 2.360 |
| 26 | 7 | 2039 | 2.357 |
| 33 | 10 | 2039 | 2.353 |
| 28 | 7 | 2038 | 2.342 |
| 34 | 10 | 2038 | 2.341 |
| 35 | 10 | 2038 | 2.340 |

Note: Strategies organized in order of decreasing age-standardized incidence rate within group. Heat map formatting for age-standardized incidence rate shows lower cervical cancer incidence rates in green and higher rates in red compared to the rate estimated under existing prevention policies, assuming 25% reduction in vaccine efficacy for cohorts aged 24 years and older (2.330 cases per 100,000 women in year 2050).

### Table S6. Change in lifetime cancer risk compared to status-quo for women by selected cohort age (25, 30, 35) in 2023, assuming 25% reduction in vaccine efficacy for specified cohorts

| **Scenario** | **Change in lifetime cancer risk*** | | |
| --- | --- | --- | --- |
|  | **25** | **30** | **35** |
| Vax up to age 25, 10-yearly screening | 10.5% | 6.5% | 7.0% |
| Vax up to age 25, 7-yearly screening | 2.1% | 2.4% | 2.8% |
| Vax up to age 25, 5-yearly screening | -2.7% | 0.0% | 0.0% |
| Vax up to age 30, 10-yearly screening | 7.3% | 0.3% | 5.6% |
| Vax up to age 30, 7-yearly screening | -0.1% | -3.6% | 1.6% |
| Vax up to age 30, 5-yearly screening | -4.9% | -5.1% | -0.8% |
| Vax up to age 35, 10-yearly screening | 6.1% | -1.9% | 0.8% |
| Vax up to age 35, 7-yearly screening | -1.2% | -5.4% | -2.8% |
| Vax up to age 35, 5-yearly screening | -5.9% | -6.6% | -5.1% |

* Positive value indicates increase in lifetime cancer risk and negative value indicates decrease in lifetime cancer risk compared to status-quo.

Note: Vax = vaccinate incrementally to maximum achieved vaccination coverage for previously vaccinated cohorts (females aged 12–32 and males aged 12–17 in 2023) and to 20% coverage for vaccine-naïve cohorts (females aged 33–35 and males aged 18–35 in 2023).

### Table S7. Elimination timeframe and age-standardized incidence rate associated with ‘EVEN FASTER’ scenarios in Norway, assuming the population structure of the World Female Population 2015 for age-standardization

| Vaccinate at maximum coverage up to specified age in 2023 | Screening frequency (years) | Year elimination (<4 cases per 100,000 women) achieved | Age-standardized incidence rate (cases per 100,000 women) in year 2050 |
| --- | --- | --- | --- |
| Status-Quo | 5 | 2034 | 1.723 |
| **Group 1. Vaccination-only strategies that decreased age-standardized incidence rate in year 2050 but did not accelerate cervical cancer elimination compared to status-quo** | | | |
| 25 | 5 | 2034 | 1.651 |
| 26 | 5 | 2034 | 1.639 |
| 27 | 5 | 2034 | 1.625 |
| 28 | 5 | 2034 | 1.609 |
| 29 | 5 | 2034 | 1.599 |
| 30 | 5 | 2034 | 1.583 |
| 31 | 5 | 2034 | 1.569 |
| 32 | 5 | 2034 | 1.557 |
| 33 | 5 | 2034 | 1.542 |
| 34 | 5 | 2034 | 1.542 |
| 35 | 5 | 2034 | 1.538 |
| **Group 2. Vaccination-only strategies that decreased age-standardized incidence rate in year 2050 and accelerated cervical cancer elimination compared to status-quo** | | | |
| **--** | **--** | **--** | **--** |
| **Group 3. Strategies that de-intensified screening and decreased age-standardized incidence rate in year 2050, but did not accelerate cervical cancer elimination compared to status-quo** | | | |
| 34 | 10 | 2034 | 1.720 |
| 28 | 7 | 2034 | 1.709 |
| 35 | 10 | 2034 | 1.707 |
| 29 | 7 | 2034 | 1.696 |
| 30 | 7 | 2034 | 1.680 |
| 31 | 7 | 2034 | 1.677 |
| 33 | 7 | 2034 | 1.655 |
| 32 | 7 | 2034 | 1.654 |
| 34 | 7 | 2034 | 1.642 |
| 35 | 7 | 2034 | 1.627 |
| **Group 4. Strategies that de-intensified screening, decreased age-standardized incidence rate in year 2050, and accelerated cervical cancer elimination compared to status-quo compared to status-quo** | | | |
| **--** | **--** | **--** | **--** |
| **Group 5. Strategies that de-intensified screening, did not decrease age-standardized incidence rate in year 2050, and did not accelerate cervical cancer elimination compared to status-quo** | | | |
| 25 | 10 | 2035 | 1.815 |
| 26 | 10 | 2035 | 1.806 |
| 27 | 10 | 2034 | 1.791 |
| 28 | 10 | 2034 | 1.779 |
| 29 | 10 | 2034 | 1.766 |
| 30 | 10 | 2034 | 1.755 |
| 31 | 10 | 2034 | 1.752 |
| 25 | 7 | 2034 | 1.740 |
| 26 | 7 | 2034 | 1.732 |
| 32 | 10 | 2034 | 1.730 |
| 33 | 10 | 2034 | 1.726 |
| 27 | 7 | 2034 | 1.723 |

Note: Strategies organized in order of decreasing age-standardized incidence rate within group. Heat map formatting for age-standardized incidence rate shows lower cervical cancer incidence rates in green and higher rates in red compared to the rate estimated under existing prevention policies (1.723 cases per 100,000 women in year 2050).
